# Localized network damage related to white matter hyperintensities is linked to worse outcome after severe stroke

**DOI:** 10.1101/2024.03.22.24304754

**Authors:** Samuel C Olszówka, Benedikt M Frey, Lukas Frontzkowski, Paweł P Wróbel, Winifried Backhaus, Focko L Higgen, Hanna Braaß, Silke Wolf, Chi-un Choe, Marlene Bönstrup, Bastian Cheng, Götz Thomalla, Philipp J Koch, Fanny Quandt, Christian Gerloff, Robert Schulz

**Author notes:** **Corresponding author:** Robert Schulz; University Medical Center Hamburg-Eppendorf; Martinistr. 52, 20246 Hamburg, Germany. These authors contributed equally to this work (Samuel C Olszówka, Benedikt M Frey).

## Abstract

White matter hyperintensities of presumed vascular origin (WMH), a manifestation of cerebral small vessel disease, are associated with various clinical sequelae. In stroke patients, total WMH burden is linked to recurrent cerebrovascular events and worse clinical outcome. As WMH also affect the integrity of structural large-scale brain networks, we hypothesize that the extent of WMH-related network damage carries relevant information to explain outcome variability in addition to global WMH volume. Clinical and structural brain imaging data of 33 severely affected acute stroke patients were analysed from two independent cohorts. Imaging data were acquired within the first two weeks after stroke. WMH-related localized and global network damage was derived, involving cortical and subcortical brain regions of an extended motor network. Using ordinal logistic regression analyses, network damage was associated with functional outcome, operationalized by the modified Rankin Scale, at follow-up after three to six months. WMH were linked to a significant disconnection of multiple ipsilesional and contralesional cortical and subcortical brain regions. Global as well as localized periventricular WMH-related network damage affecting distinct brain regions of both hemispheres, including the precentral and the inferior frontal gyrus, areas of the dorsolateral prefrontal cortex, the insula, and multiple subcortical nuclei, was independently associated with a worse outcome after adjustment for baseline symptom burden, age, brain infarct volume and total WMH volume. Total and deep WMH-related network disturbances did not show similar associations. This study shows that periventricular WMH-related network damage affecting specific brain regions of the frontal and insular lobe, and subcortical nuclei is linked to functional outcome in acute stroke patients. This supports the evolving concept of structural brain reserve and underscores the potential significance of pre-existing WMH-related network damage as a crucial factor in comprehending outcome variability after severe stroke.

## Introduction

White matter hyperintensities of presumed vascular origin (WMH) are a manifestation of cerebral small vessel disease (cSVD) and are associated with various clinical sequelae such as cognitive decline, dementia, depression as well as disturbances of gait and balance. ^1-3^ Based on their distance to the inner ventricle system, WMH can be divided into deep WMH (dWMH) and periventricular WMH (pWMH), with the two forms being related to different risk factors and neurological impairment. ^4^ WMH are also a common finding in magnetic resonance imaging (MRI) of ischemic stroke patients, and 25% of ischemic strokes can be attributed to the presence of cSVD. ^5^ WMH burden is linked to higher probabilities of first-ever^3^ or recurrent strokes, ^6^ as well as a worse outcome after stroke. ^6-8^

Currently, the factors driving the association between WMH and outcome after stroke remain largely unknown. From a clinical perspective, WMH might simply reflect the systemic burden of vascular risk factors, even though the association of WMH and clinical outcome remained stable in adjusted meta-analyses. ^6,7^ WMH may also exert a behavioural influence onto stroke recovery, for instance through cognitive dysfunctions related to WMH which reduce patients’ adherence to and participation in neurorehabilitative treatment. ^7^ However, from a systems neuroscience perspective, WMH may impact recovery trajectories by affecting large-scale structural and functional brain networks^9,10^ as numerous studies have consistently reported that the state of structural brain networks, quantified early after acute stroke, can inform about subsequent recovery. ^11-13^ WMH-related network damage could serve as a surrogate for pre-existing disturbances in brain networks, and it could be a novel approach to better understand the underlying mechanisms of brain reserve capacity^14^ and outcome variability after stroke, as recently evidenced for cortical and cerebellar brain anatomy. ^15,16^

The present study sought to answer the question whether the extent of WMH-related network damage, quantified early after severe ischemic stroke, can explain outcome variability. To this end, clinical and structural brain imaging data of 33 severely affected acute stroke patients were analysed. ^17,18^ WMH-related localized and global network damage was calculated for multiple cortical and subcortical brain regions of an extended motor network. Network damage was associated with functional outcome, operationalized by the modified Rankin Scale (mRS) at follow-up.

## Methods

### Participants

The data used in this study were obtained by combining two independent cohorts of acute stroke patients whose details have been published previously (study A: n=30^17^ and study B: n=61^18^). We already used this approach in two previous studies. ^15,16^ All patients were treated at the University Medical Center Hamburg-Eppendorf between 2012 and 2020. The final sample size comprised 33 datasets. Please see the Supplementary Methods for details of cohort integration, a flowchart of dataset composition is given in Supplementary Figure 1. Patients received structural MRI of the brain within 3-14 days after stroke onset (3-14 days in study A and 3-5 days in study B). In three patients, imaging including T1- and T2-weighted data was available at a later point (n=1 within 1 month and n=2 within 3 months). In one patient, FLAIR data was not available, and T2* data was used for further analysis. Initial deficit was assessed by the National Institute of Health Stroke Scale (NIHSS) at the time of study inclusion (i.e. after acute treatment). Functional outcome was operationalized by the mRS in the late subacute stage of recovery three months after stroke, or after six months in four patients in which data for the earlier time point were not available. The original studies were conducted in line with the ethical declaration of Helsinki and were granted permission by the local ethics committee of the Chamber of Physicians Hamburg. All participants or their legal guardian provided informed consent.

### Image acquisition and processing

Imaging was performed with the same 3 Tesla Skyra MRI scanner (Siemens Healthineers, Erlangen, Germany). Details on sequences and image processing are given in the Supplementary Methods. The distribution of stroke and WMH lesions is illustrated in Supplementary Figure 2. Normalized masks of WMH, pWMH, and dWMH were analyzed via the NeMo tool (v2.1a8) ^19^ (https://kuceyeski-wcm-web.s3.us-east-1.amazonaws.com/upload.html). This tool allows the projection of binary masks in MNI space over a healthy brain network, averaged from whole-brain tractography data of 420 healthy individuals derived from the Human Connectome Project, and calculates the resulting change of connectivity (ChaCo, i.e., loss of connectivity) in a highly reproducible way. ^19^ Details are given in the Supplementary Material. As mRS is an outcome measure dominated by stroke deficits in the motor domain, ^20^ an extended motor network atlas was chosen as the output resolution for the NeMo analyses which encompasses 106 cortical and subcortical brain regions from the brainnetome atlas structurally connected to the upper limb representation of the primary motor cortex. ^21^ An illustration of this individual atlas is given in Supplementary Figure 3. Figure 1 illustrates the steps of image processing and disconnectivity measure acquisition.

**Figure 1.**
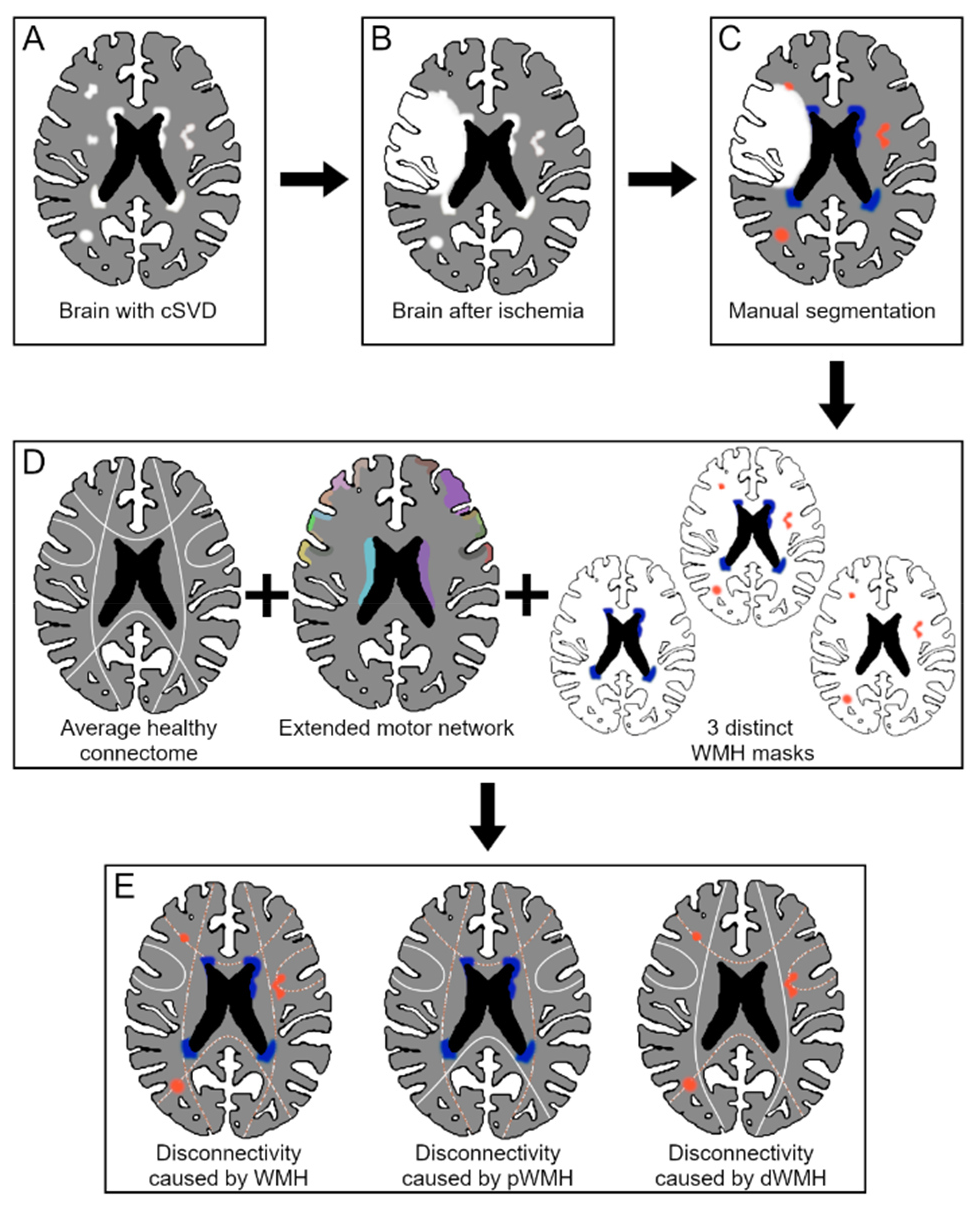
Image processing and disconnectivity measures of WMH-related networks. Overview of image processing and disconnectivity measure acquisition. **A**: Visible WMH appearing in T2-weighted MRI due to cerebral small vessel disease. **B**: Stroke-lesion which overlays the pre-existing WMH. **C**: Manual WMH segmentation. Some are hidden by the stroke-lesion and are therefore not included in the segmentation. **D**: Segmented WMH are used to create three binary masks: one mask containing all WMH, one mask containing dWMH only, one mask containing pWMH only. These three masks are separately processed by the Network Modification (NeMo) Tool with an extended motor network serving as the reference connectome. **E**: The output of the NeMo analyses provides estimates of localized network damage, i.e., disconnection due to WMH, affecting specific cortical and subcortical regions.

### Statistical Analysis

Statistical analysis was performed using R version 4.3.1. Unpaired one-sided *t*-tests were conducted to assesses which regions were significantly disconnected from the remaining brain network at group level. Afterwards, cortical, and subcortical regions with a strongly skewed distribution of those values (γ1 > 1.3 or < -1.3) or a median of 0 or 1, respectively, were excluded from further analyses. The final set of regions undergoing statistical modelling is given in Supplementary Table 1. Regional ChaCo values, i.e., the extent of network damage affecting specific regions, were binarized into “low” and “high” disconnectivity based on median split with dichotomization. Odd number of participants (n=33) resulted in a true median, which was assigned to the “small” disconnectivity group.

Ordinal logistic regression models were fitted for mRS at follow-up as the dependent variable. Global and region-specific WMH-related network disconnectivity (high or low) was treated as the independent variable of interest. The following widely established nuisance variables were primarily included to adjust target effects: age, NIHSS at study inclusion, stroke lesion volume, and total WMH volume. Odds ratios (OR), including 95% confidence intervals (CI), and *P* values were extracted from the models. An OR of higher than 1 would indicate a higher probability of scoring one level higher in mRS at follow-up, i.e., having a worse functional outcome. To assess the amount of additionally explained variance by the extent of network damage affecting each region (predictor of interest), gain in R^2^ was given when compared to the comparative base model without the predictor of interest. Base model statistics are summarized in Supplementary Table 2. Analyses were repeated for each region for WMH and the subcomponents dWMH and pWMH in separate iterations according to Supplementary Table 1. After ensuring the robustness with a leave-one-patient-out analysis (LOOA), model-derived *P* values were corrected for multiple testing using the false discovery rate (FDR) correction. Please see the Supplementary Material for details of the statistical analysis. All findings were reported in accordance with the STROBE reporting guideline (Supplementary Material).^22^

## Data availability

Data will be made available by the authors upon reasonable request.

## Results

### Demographical and clinical data

Table 1 summarizes demographical and clinical data of the 33 severely affected first-ever ischemic stroke patients included in this analysis.

**Table 1.** Demographical and clinical data. Abbreviations: mRS, modified Rankin Scale; NIHSS, National Institute of Health Stroke Scale; WMH, White Matter Hyperintensities of presumed vascular origin; pWMH, periventricular WMH; dWMH, deep WMH

| Clinical characteristic |  | Total (n=33) |
| --- | --- | --- |
| Age, years (mean, SD) |  | 70.7 (12.2) |
| Sex |  |  |
| - | Male (%) | 16 (48%) |
| - | Female (%) | 17 (52%) |
| Affected hemisphere |  |  |
| - | Left (%) | 13 (39%) |
| - | Right (%) | 20 (61%) |
| Handedness |  |  |
| - | Left (%) | 2 (6%) |
| - | Right (%) | 31 (94%) |
| Dominant side stroke (%) |  | 13 (39%) |
| Brainstem stroke (%) |  | 5 (15%) |
| Initial therapy |  |  |
| - | Thrombolysis only (%) | 5 (15%) |
| - | Thrombectomy only (%) | 1 (3%) |
| - | Both (%) | 13 (39%) |
| NIHSS at study inclusion |  |  |
| - | Mean (SD) | 7.8 (3.6) |
| - | Range | 1-17 |
| mRS at follow-up |  |  |
| - | Median (Q1-Q3) | 3 (1-4) |
| - | Range | 1-4 |
| Stroke volume (cm<sup>3</sup>, mean, SD) |  | 37.9 (50.4) |
| WMH volume (cm<sup>3</sup>, mean, SD) |  | 15.3 (10.6) |
| pWMH volume (cm<sup>3</sup>, mean, SD) |  | 9.5 (6.3) |
| dWMH volume (cm<sup>3</sup>, mean, SD) |  | 5.8 (4.7) |

### WMH-related network damage after stroke

Out of the 106 regions of the extended motor network, 100 regions were significantly disconnected by total WMH, 100 by pWMH and 98 by dWMH. A cortical projection of significant mean ChaCo values is given in Figure 2 A-C. Across cortical regions exhibiting significant disconnection, the average ChaCo value was 0.19 ± 0.06 (mean ± SD) for total WMH, 0.09 ± 0.05 for pWMH, and 0.13 ± 0.04 for dWMH. Deep WMH led to higher levels of cortical disconnectivity than pWMH (*t*(67) = 5.2, *P*<0.001) despite that dWMH volume was lower than pWMH volume (Table 1, *t*(32) = -6.6, *P*<0.001). Across subcortical regions exhibiting significant disconnection, the average ChaCo value was 0.20 ± 0.09 for total WMH, 0.15 ± 0.07 for pWMH, and 0.09 ± 0.06 for dWMH. Periventricular WMH led to higher levels of subcortical disconnectivity than dWMH (*t*(29) = 3.9, *P*<0.001) in accordance to pWMH volume being higher highest mean disconnectivity (i.e. ChaCo) caused by total WMH were dorsal caudate (0.39 ± 0.16, Mean ± SD), dorsolateral putamen (0.35 ±0.22), sensory (0.33 ± 0.24) and posterior parietal thalamus (0.31 ± 0.19) as well as lateral area 10 (0.30 ± 0.13) on the contralesional hemisphere. Areas with highest mean disconnectivity caused by pWMH were contralesional (0.36 ± 0.16) and ipsilesional (0.27 ± 0.20) dorsal caudate, as well as posterior parietal (0.25 ± 0.17) and sensory thalamus (0.25 ± 0.21) on the contralesional hemisphere. Highest mean disconnectivity caused by dWMH affected contralesional dorsolateral putamen (0.27 ± 0.20) and both contralesional (0.25 ± 0.22) and ipsilesional (0.25 ± 0.21) rostrodorsal area 39.

### Association between disconnectivity measures and functional outcome

The disconnectivity of multiple regions affected by pWMH was significantly linked to a worse outcome at follow-up, independent from total WMH volume, age, lesion volume and initial NIHSS. Specifically, on the ipsilesional hemisphere, areas of the middle frontal gyrus contributing to the dorsolateral prefrontal cortex (DLPFC) including ventral Brodmann area 9/46, area 46, ventrolateral area 8, lateral area 10 and inferior frontal junction additionally explained up to 17.0% of variance. Patients exhibiting higher network disconnectivity in these regions had higher odds of showing higher mRS at follow-up when compared to patients with lower network disconnectivity (Table 2, Figure 2). On the inferior frontal gyrus, higher disconnection of ventral area 44, the inferior frontal sulcus, as well as insular gyrus, were similarly associated with worse outcome after stroke. On the contralesional hemisphere, associations between pWMH-related network damage and worse outcome were detected for upper limb area 4 of the primary motor cortex and caudal dorsolateral area 6 of the precentral gyrus, area 9/46 and inferior frontal gyrus of the middle and inferior frontal gyrus, respectively. Finally, multiple contralesional subcortical nuclei, the amygdala, globus pallidus, hippocampus, and thalamus did also show significant associations. Compared to pWMH, localized network disconnectivity of WMH or dWMH did not show similar relationships with outcome after stroke (all *P*_FDR_>0.1).

**Table 2.**
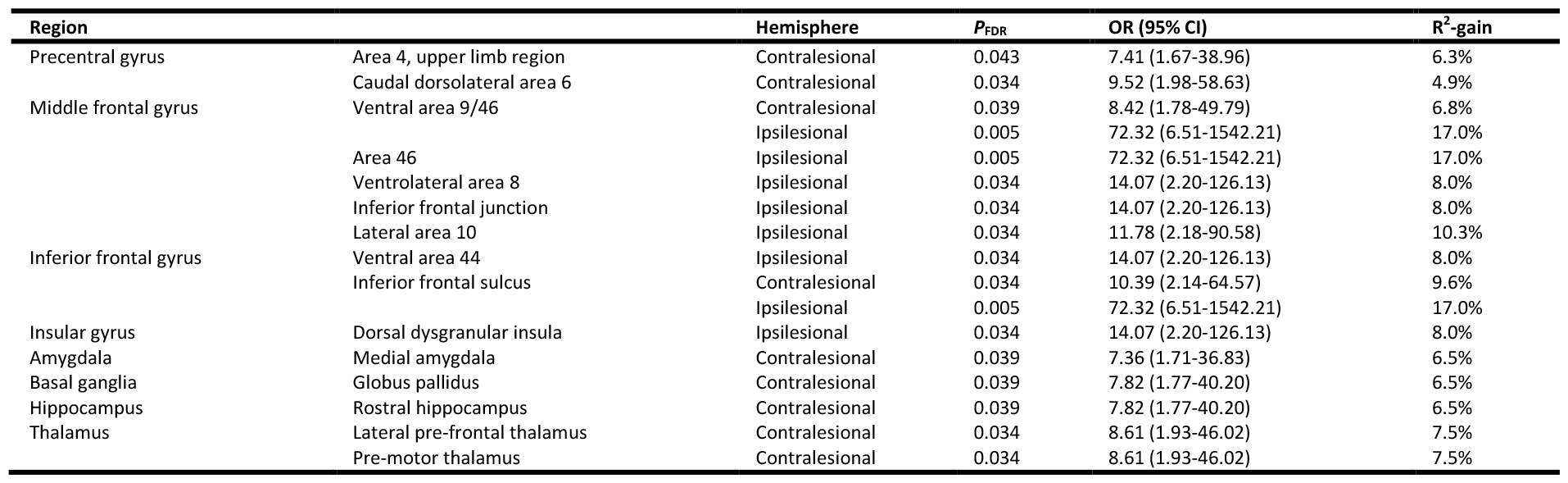
Localized pWMH-related network damage relates to outcome after stroke. Significant associations between dichotomized pWMH-related network damage affecting cortical and subcortical brain regions and outcome after stroke. Results are adjusted for age, initial NIHSS, lesion and total WMH volumes. ORs with 95% CIs are given for patients with higher disconnectivity (reference) of rising one level in mRS compared to patients with lower disconnectivity for the specific region. *P* values are FDR corrected (*P*_FDR_) for 93 tests. R^2^-gain is given as additional explained variance (R^2^) compared to R^2^ of the base model (39.5%). All results remain stable after LOOA with *P*<0.05 (uncorrected).

**Figure 2:**
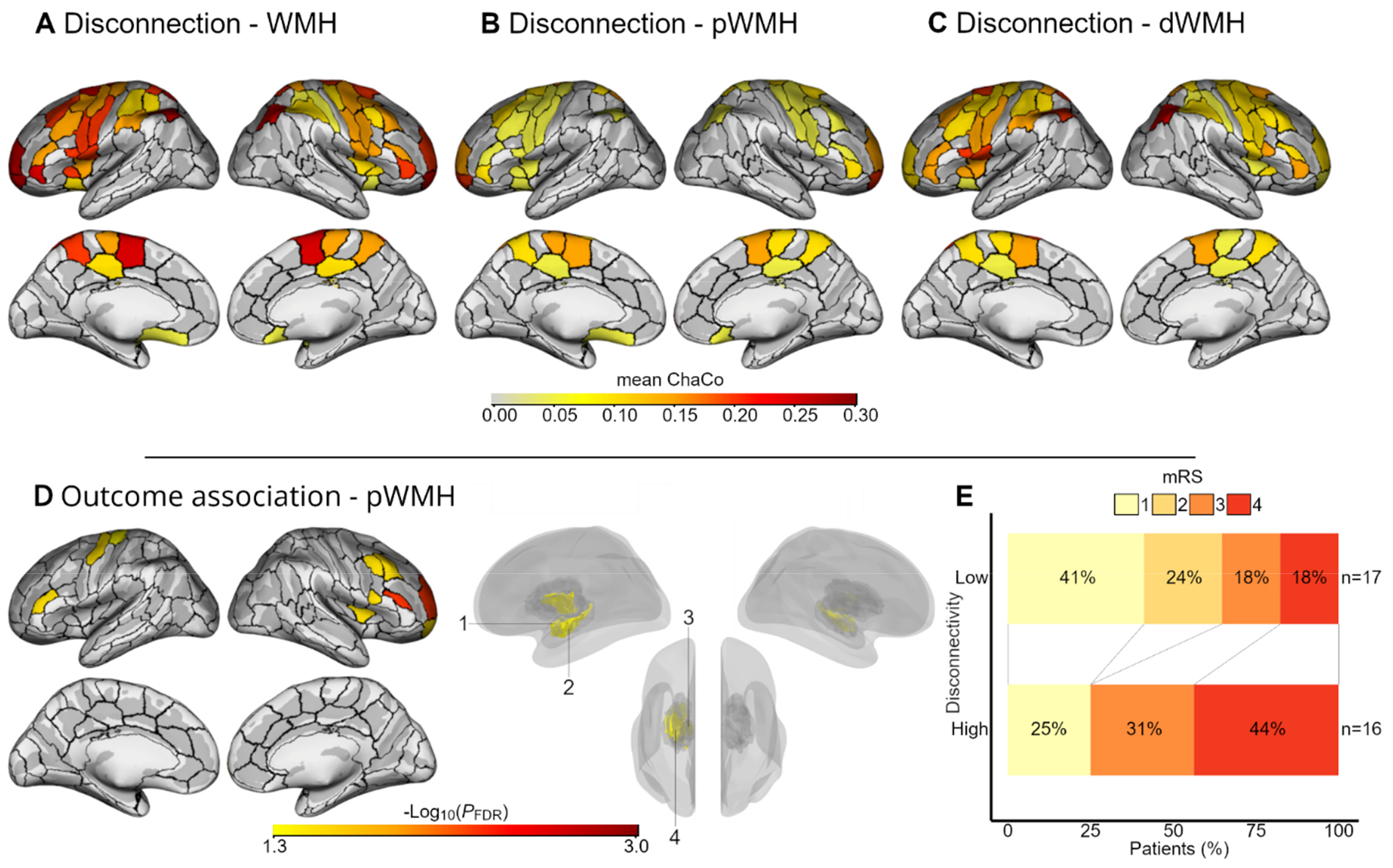
Localized WMH-related network disconnections and correlations with outcome after stroke. **A-C**: Significant region wise mean ChaCo values, presented separately for total (A), periventricular (B, pWMH) and deep WMH (C, dWMH), visualized as a brain surface overlay. ChaCo value of 1 correspond to total disconnection from the remaining brain network, 0 corresponds to no disconnection at all. Unpaired one-tailed *t*-test against 0 was performed to assess significance of these measures. **D**: Correlation between dichotomized disconnection of brain regions, caused by pWMH, and outcome after stroke with *P*_FDR_-values (-Log_10_ transformed) of associations projected onto the corresponding cortical and subcortical areas. 1: Medial amygdala. 2: Rostral hippocampus. 3: Lateral pre-frontal and pre-motor thalamus. 4: Globus pallidus. **E**: Shift-Plot depicting influence of “high” vs. “low” disconnectivity group on mRS at follow-up, based on mean disconnectivity of all significant regions (n=17, see Table 2). For illustration purposes only.

For sensitivity analyses, we recomputed the base model considering pWMH and dWMH volumes, and, for a network perspective, also the average ChaCo for WMH, pWMH, and dWMH instead of total WMH volume, respectively. In these alternative base models, only pWMH-mediated global network damage showed a significant contribution to the base model (*P*=0.025, Supplementary Tables 3 and 4) and improved the explanation of variance from 39.5% to 45.4%. Primary outcome models based on the subcomponent of pWMH (n=93) were refit adjusted for global pWMH network damage instead of WMH volume. Results remained largely stable after using this improved base model (Supplementary Table 5).

## Discussion

The main finding of the present work was that localized pWMH-related network damage disconnecting distinct brain regions from an extended motor network was linked to worse outcome after a severe stroke. The association was independent of the initial deficit, age, overall volumes of WMH and the stroke lesion. Specifically, we showed that areas contributing to the ipsilesional DLPFC, insular cortex, contralesional primary and secondary motor areas as well as subcortical nuclei including the thalamus exerted a negative influence on stroke recovery when disconnected from large-scale networks by pWMH. Herewith, this study adds a novel network perspective to previous data of global WMH burden as a risk factor for worse outcome after stroke.

WMH burden has repeatedly been associated with worse outcome after stroke in clinical populations. ^6,7^ However, in patients with severe deficits, caused by large vessel occlusion, previous studies have questioned such observations as they did not find a clear relationship between WMH and revascularizing treatment gains and outcome. ^23,24^ In agreement with these data, we did not detect significant correlations between the established WMH volume and mRS at follow-up in our cohort, neither for total WMH, nor for pWMH and dWMH (Supplementary Tables 2 and 3). In contrast, we found that the degree of network disconnectivity affecting specific cortical and subcortical brain regions associated with pWMH was correlated with functional outcome. Interestingly, already substituting the WMH volume in the base model by pWMH-related global network damage explained an additional amount of variance in mRS at follow-up which highlights that WMH-related network damage seems to be particularly informative for outcome inference after severe stroke.

We found only pWMH-related global and localized network damage to be linked to clinical outcome, but not network damage derived from total WMH or dWMH. A similar observation was made for recovery after primary intracerebral haemorrhage where higher pWMH, but not dWMH volumes were associated to an unfavourable long-term outcome. ^25^ Another study could link the extent of cortical atrophy to pWMH-related disruption of connecting white matter tracts, an association which was not evident for total WMH or dWMH-affected networks. ^26^ There are different possible explanations for these differences. On the one hand, it is possible that brain regions involved in recovery processes are simply connected by fibres running through regions typically affected by pWMH. On the other hand, we speculate statistically that the relatively stable spatial distribution of pWMH could result in more robust effects at group-level in smaller cohorts when compared to dWMH.

The region-specific assessment of network damage related to pWMH allowed us to answer whether localized network disconnection affecting specific cortical and subcortical regions might impact clinical outcome. Four regions will be discussed in the following.

On the ipsilesional hemisphere higher disconnection of areas contributing to the DLPFC was correlated with a worse outcome. Instead of short association fibres, whose disruption would expectedly be mostly driven by dWMH, ^26^ pWMH might act on the DLPFC through disruption of thalamic association fibres. ^27^ This is further supported by the concomitant significant disconnection of thalamic brain regions (Table 2, Figure 2) and findings of a previous study showing that WMH are to a large extent located within the anterior thalamic radiatio and superior longitudinal fasciculus, ^28^ which both connect to the DLPFC.

Previous studies have shown that non-invasive stimulation of the DLPFC might serve as one innovative treatment approach to enhance functional recovery after stroke, particularly in the cognitive domain.^29^ Herein, stimulation effects were more likely to affect memory, attentional and executive^30^ than motor functions. ^31^ One study found that an increased functional connectivity between contralesional DLPFC and mid-ventrolateral prefrontal cortex was linked to better hand function in chronic stroke patients. ^32^ Vice versa, others reported that subacute stroke patients, in which ipsilesional DLPFC areas gradually disconnected from larger connectomes by the lesion, were at higher risks for impaired natural recovery. ^11^ The present study furthers these insights and shows that DLPFC disconnection from larger brain networks, potentially also caused by pre-existing pWMH, can increase, at least partly, the risk for an unfavourable outcome.

A second significant association for pWMH-related network damage and outcome was found for contralesional precentral gyrus including the primary motor cortex and areas contributing to the dorsolateral premotor cortex and ventral premotor cortex. Given the associations between pWMH-related disconnectivity and cortical atrophy previously described, ^26^ we noticed an interesting parallel comparing the current results to an earlier report from our group. Based on a similar cohort of severely impaired stroke patients, we found that reduced cortical thickness of the contralesional precentral gyrus at baseline was significantly associated with a worse outcome. ^16^ Hence, the relationship of clinical outcome and cortical thickness might be driven by pWMH-related network disturbances over time. Combined analyses of baseline cortical anatomy and WMH disconnections are warranted to explore the interrelationship between both measures and their importance for recovery systematically, following the emerging concepts of structural brain reserve after stroke. ^14^

Finally, the dorsal insula on the ipsilesional side and subcortical nuclei on the contralesional side, including the pre-motor and lateral pre-frontal thalamus, were identified as risk factors for a poorer outcome when disconnected from larger brain networks by pWMH. These findings are in line with prior research. For instance, lesions within the insular lobe have been associated with larger initial deficits, ^33^ worse outcome, ^34^ and higher mortality. ^35^ For the thalamus, previous reports have linked ipsilesional thalamic volume and secondary thalamic atrophy after stroke to worse sensorimotor^36^ and cognitive performance. ^37^ WMH-related network damage affecting the insula or thalamus might make such highly interconnected hubs particularly susceptible to further, and then, critical disconnection caused by the stroke lesion itself. Threatened by disconnection and secondary atrophy, key nodes of the human motor system such as the thalamus, could lose important regulatory functions necessary for normal movements^38^ or, as animal studies have shown, re-learning processes. ^39^

## Limitations

There are several important limitations worth noting. First, analyses were conducted on a small sample of acute stroke patients. However, statistical modelling was corrected for multiple testing and a LOOA analysis was added to enhance the robustness of the findings. However, sensitivity and generalizability of the present analyses are likely to be reduced, and presented OR must be interpreted with care. Therefore, prospective studies are needed to reaffirm reported results. Second, brain imaging for acute lesion volume and WMH segmentation, based on T1- and T2-weighted MRI, was conducted after stroke, mostly within the first two weeks after stroke but up to three months after stroke in two patients. Hence, atrophy processes influencing lesion topography and segmentation, image normalization, and altering WMH with a small risk for additional de novo manifestations of WMH after stroke may influence the present modelling results. For further sensitivity analyses, we repeated all analyses while excluding three patients whose imaging was later than two weeks. Most top performing regions remained stable (Supplementary Table 6). This result, however, must be interpreted with caution in view of the low precision of the OR estimates. Third, WMH located within the primary stroke lesion could not be properly delineated, their extent remained elusive, and they were not included in the binary WMH masks. Also, WMH volumes were therefore underestimated with a bias towards patients with larger stroke lesions. We decided not to address this problem, e.g., by mirroring WMH distributions from the contralesional to the ipsilesional hemisphere to estimate a *pre-stroke* state of the lesioned brain, as such an approach is neither straightforward nor free of further limitations. Fourth, the connectome analysis was based on an arbitrary value of complete streamline disruption evoked by all individual WMH lesions. Whether a more fine-grained definition of the disruptive impact of each WMH lesion, e.g., based on quantitative T2-weighted data or diffusion-based microstructural information, might alter the present findings remains an interesting research question for future multimodal work. Fifth, mRS was used as the outcome measure, a clinical score of global disability, dominated by preserved functions of the motor domain. Therefore, the reference network for connectome analyses was biased towards an extended motor network. ^21^ Alternative outcome measures in the cognitive or language domains might call for alternative reference networks and alter the present results.

## Conclusion and outlook

This study shows that pWMH disconnect specific brain regions including primary and premotor cortices, dorsolateral prefrontal cortices, the insula, and various subcortical nuclei such as the thalamus from larger brain networks. Severely impaired stroke patients with higher amount of disconnection in these key areas of cognitive, learning and motor functioning are at an increased risk of a worse outcome, independent from the initial deficits, age, lesion volume and total WMH volume. Hence, this work adds novel network, and potentially also mechanistic, data to previous research promoting WMH as an important risk factor for impaired recovery after stroke. Regarding clinical applications it may lead to interesting new research questions. For instance, could patients with network critical WMH distributions benefit from intensive adjustment of cardiovascular risk factors more than patients with less critical network damage? Or could WMH-related network damage with consecutive disruption of cognitively important brain regions help to stratify patients towards motor and cognitive neurorehabilitative training or non-invasive brain stimulation of non-motor areas to enhance recovery after stroke?

## Acknowledgments

None.

## Sources of Funding

This work was funded by the Deutsche Forschungsgemeinschaft (DFG, German Research Foundation) SFB 936 - 178316478 – projects C1 to C.G., C2 to G.T. and Deutsche Forschungsgemeinschaft (DFG, German Research Foundation) and the National Science Foundation of China (NSFC) in project Crossmodal Learning, TRR-169/A3 to F.H. and C.G., and the Else Kröner-Fresenius-Stiftung (2016_A214 to R.S.). R.S. and C.U.C. are supported by an Else Kröner Exzellenzstipendium from the Else Kröner-Fresenius-Stiftung (2020_EKES.16 to R.S., 2018_EKES.04 to C.U.C.). F.Q. is supported by the Gemeinnützige Hertie-Stiftung (Hertie Network of Excellence in Clinical Neuroscience).

## Disclosures

None.

## Notes

### Competing Interest Statement

The authors have declared no competing interest.

### Author Declarations

The original studies were conducted in line with the ethical declaration of Helsinki and were granted permission by the local ethics committee of the Chamber of Physicians Hamburg. All participants or their legal guardian provided informed consent.

