## Supplemental Material for "Localized network damage related to white matter hyperintensities is linked to worse outcome after severe stroke"

[1] University Medical Center Hamburg-Eppendorf, Department of Neurology, Hamburg, Germany
[2] University Medical Center Frankfurt, Department of Neurology, Frankfurt, Germany
[3] Department of Neurology, University Hospital Schleswig-Holstein, Campus Lübeck, Germany
[†] These authors contributed equally to this work

### Supplementary Methods

#### Cohort integration

For cohort integration, inclusion criteria from study A were retrospectively applied for study B, in line with previous reports,[15,16] that were modified Rankin Scale (mRS) > 3 or a Barthel Index ≤ 30 at admission, age ≥ 18 years, first-ever ischemic stroke with persistent motor deficit of the upper limb and no prior history of severe psychiatric or neurological disorders. After excluding incomplete (n=28) or qualitatively insufficient MRI images (n=4), the final sample size comprised 33 datasets.

#### Image acquisition and processing

Imaging was performed with the same 3 Tesla Skyra MRI scanner (Siemens Healthineers, Erlangen, Germany) in all included patients using a 32-channel head coil and a T1-weighted magnetization-prepared rapid gradient echo sequence (repetition time (TR) = 2500 ms, echo time (TE) = 2.12 ms, flip angle = 9°, number of coronal slices = 256, voxel size = 0.8 × 0.8 × 0.9 $mm^3$ and field of view (FOV) = 240 mm) as well as T2-weighted fluid attenuated inverse recovery (FLAIR) datasets (TR = 9000 ms, TE = 86 ms, time to inversion = 2500 ms, flip angle = 150°, number of transversal slices = 43, voxel size = 0.7 × 0.7 × 3.0 $mm^3$ and FOV = 230 mm).

WMH and stroke lesions were delineated manually by a trained rater on FLAIR scans using the "Active Contour Segmentation" tool, provided by ITK-SNAP (v3.8.0).[40] To estimate intra-rater-reliability, the segmentation process was repeated in eight randomly selected patients with good results (94.9% for WMH volumes and 98.7% for stroke lesion volumes). Masks of WMH and stroke lesions were normalized to the Montreal Neurological Institute (MNI) 152 standard space using the Advanced Normalization Tools.[41] Normalized WMH masks were divided into pWMH and dWMH by a 10 mm distance threshold to the ventricles.[4] The quality of the image processing steps was carefully assured by repeated visual inspection.

The parcellation was dilated by 1 mm and region wise disconnectivity was calculated using an output resolution of 1 mm and a probabilistic tractography algorithm. Resulting change of connectivity values (ChaCo values), ranging from 0 (not disconnected from the extended motor network) to 1 (fully disconnected) were processed using Python version 3.10 (https://github.com/kjamison/nemo). Disconnectivity values below 0.02 were set to 0 and values above 1 were set to 1, as such values can be attributed to noise within the NeMo Tool.

#### Statistical Analysis

Unpaired one-sided *t*-tests were conducted to assesses which regions were significantly disconnected from the remaining brain network at group level ($P<0.05$), comparing each regions mean ChaCo value against 0. Mean ChaCo of subcortical and cortical regions was calculated separately, and difference in these measures between each of the three groups (total WMH, pWMH, dWMH) was determined using paired two-tailed *t*-tests.

Ordinal logistic regression models were fitted for mRS at follow-up as the dependent variable. Global and region-specific WMH-related high/low network disconnectivity was treated as the independent variable of interest. The following nuisance variables were included to adjust target effects: age, NIHSS at study inclusion, stroke lesion volume, and total WMH volume. Stroke lesion and WMH volumes were $Log_{10}$ transformed to improve data distribution. Given strong correlations between age and WMH,[2] and assuming a priori a correlation between WMH volume and WMH-related network damage, age and total WMH volume were included after residualization against the region-specific WMH-related disconnectivity.[15,16]

Leave-one-patient-out model analyses (LOOA) at a cut-off of $P>0.05$ were conducted to ensure robustness of the findings. False-discovery rate (FDR) correction of the model-derived *P* values of the predictor of interest was performed separately for 93 (pWMH), 91 (total WMH) and 71 (dWMH) tests within each of the three iterations, respectively.

**Supplementary Tables**

| Total WMH (WMH) | | Periventricular WMH (pWMH) | | Deep WMH (dWMH) | |
|---|---|---|---|---|---|
| Contralesional | Ipsilesional | Contralesional | Ipsilesional | Contralesional | Ipsilesional |
| A1.2.3ll | A1.2.3ll | A1.2.3ll | A1.2.3ll | A1.2.3tru | A1.2.3tru |
| A1.2.3tru | A1.2.3tru | A1.2.3tru | A1.2.3tru | A1.2.3ulhf | A1.2.3ulhf |
| A1.2.3ulhf | A1.2.3ulhf | A1.2.3ulhf | - | A12.47o | - |
| A10l | A10l | A10l | A10l | - | A23c |
| A12.47o | A12.47o | A12.47o | A12.47o | A39rd | A39rd |
| - | A13 | A13 | A13 | A40rd | A40rd |
| - | A2 | A2 | - | A44v | A44v |
| A23c | A23c | A23c | A23c | A45r | A45r |
| A39rd | A39rd | A39rd | A39rd | - | A46 |
| A40rd | A40rd | A40rd | A40rd | A4hf | - |
| A44v | A44v | A44v | A44v | A4ll | A4ll |
| A45r | A45r | A45r | A45r | A4t | A4t |
| A46 | A46 | A46 | A46 | A4tl | - |
| A4hf | A4hf | A4hf | A4hf | A4ul | A4ul |
| A4ll | A4ll | A4ll | A4ll | A5m | - |
| A4t | A4t | A4t | A4t | - | A6cdl |
| A4tl | - | A4tl | A4tl | A6cvl | A6cvl |
| A4ul | A4ul | A4ul | A4ul | A6dl | A6dl |
| A5m | A5m | A5l | - | A6m | A6m |
| A6cdl | A6cdl | A5m | A5m | A6vl | - |
| A6cvl | A6cvl | A6cdl | A6cdl | A7pc | - |
| A6dl | A6dl | A6cvl | A6cvl | A7r | A7r |
| A6m | A6m | A6dl | A6dl | A8vl | A8vl |
| A6vl | A6vl | A6m | A6m | - | A9.46v |
| A7pc | A7pc | A6vl | A6vl | dCa | dCa |
| A7r | A7r | A7pc | A7pc | dIa | dIa |
| A8vl | A8vl | A7r | A7r | dId | dId |
| A9.46v | A9.46v | A8vl | A8vl | dlPu | dlPu |
| - | cHipp | A9.46v | A9.46v | GP | GP |
| dCa | dCa | - | cHipp | IFJ | - |
| dIa | dIa | dCa | dCa | IFS | IFS |
| dId | dId | dIa | dIa | lPFtha | lPFtha |
| dlPu | dlPu | dId | dId | mAmyg | - |
| GP | GP | dlPu | dlPu | mPFtha | mPFtha |
| IFJ | - | GP | GP | mPMtha | mPMtha |
| IFS | IFS | IFJ | IFJ | NAC | - |
| lPFtha | lPFtha | IFS | IFS | - | Otha |
| mAmyg | mAmyg | lPFtha | lPFtha | PPtha | PPtha |
| - | mPFtha | mAmyg | - | rHipp | - |
| mPMtha | mPMtha | - | mPFtha | Stha | Stha |
| NAC | NAC | mPMtha | mPMtha | vCa | - |
| Otha | Otha | NAC | NAC | - | vIa |
| PPtha | PPtha | Otha | Otha | vId.vIg | vId.vIg |
| rHipp | rHipp | PPtha | PPtha | vmPu | vmPu |
| Stha | Stha | rHipp | rHipp | | |
| vCa | vCa | Stha | Stha | | |
| - | vIa | vCa | vCa | | |
| vId.vIg | vId.vIg | - | vIa | | |
| vmPu | vmPu | vId.vIg | vId.vIg | | |
| | | vmPu | vmPu | | |

**Supplementary Table 1. Final list of regions undergoing statistical modeling for each of the three iterations.** Comprehensive list of all regions undergoing statistical modeling for each of the three test-regimen. After excluding regions with skewed ChaCo distribution ($\gamma_1 > 1.3$ or $< -1.3$) or median of 0 or 1, respectively, 91 regions were included for the analysis of total WMH, 93 for periventricular WMH, and 71 for deep WMH.

| Predictor | OR (95% CI) | *P* |
|---|---|---|
| NIHSS at admission | 1.53 (1.16-2.02) | <0.001 |
| Age | 1.04 (0.99-1.10) | 0.140 |
| $Log_{10}$(stroke lesion volume) | 0.99 (0.62-1.59) | 0.978 |
| $Log_{10}$(WMH volume) | 1.88 (0.70-5.08) | 0.202 |

**Supplementary Table 2. Base model details.**
Predictor of interest was mRS at follow up. Stroke lesion volume and WMH volume were $Log_{10}$-transformed to account for skewed distribution. The model was able to explain 39.5% of variance ($R^2$).

| Predictor | OR (95% CI) | *P* | Predictor | OR (95% CI) | *P* |
|---|---|---|---|---|---|
| NIHSS at admission | 1.55 (1.19-2.12) | <0.001 | NIHSS at admission | 1.52 (1.18-2.06) | <0.001 |
| Age | 1.03 (0.98-1.09) | 0.202 | Age | 1.05 (0.99-1.11) | 0.097 |
| $Log_{10}$(stroke lesion volume) | 0.99 (0.60-1.60) | 0.966 | $Log_{10}$(stroke lesion volume) | 1.00 (0.62-1.60) | 0.995 |
| $Log_{10}$(pWMH volume) | 2.51 (0.86-8.30) | 0.093 | $Log_{10}$(dWMH volume) | 1.34 (0.65-2.85) | 0.425 |

**Supplementary Table 3. Effect of pWMH and dWMH volume on outcome.**
Predictor of interest was mRS at follow up. Stroke lesion volume and pWMH volume were $Log_{10}$-transformed to account for skewed distribution. Models were able to explain 41.6% and 37.6% of variance ($R^2$) for pWMH and dWMH, respectively.

| Predictor | *P* | Predictor | *P* | Predictor | *P* |
|---|---|---|---|---|---|
| NIHSS at admission | 0.002 | NIHSS at admission | 0.002 | NIHSS at admission | 0.002 |
| Age | 0.155 | Age | 0.288 | Age | 0.136 |
| $Log_{10}$(stroke lesion volume) | 0.870 | $Log_{10}$(stroke lesion volume) | 0.745 | $Log_{10}$(stroke lesion volume) | 0.915 |
| Mean WMH ChaCo | 0.225 | Mean pWMH ChaCo | 0.025 | Mean dWMH ChaCo | 0.345 |

**Supplementary Table 4. Effect of total WMH, pWMH and dWMH mediated mean change of connectivity (ChaCo) on outcome.**
Predictor of interest was mRS at follow up. Stroke lesion volume was $Log_{10}$-transformed to account for skewed distribution. Models were able to explain 39.2%, 45.4% and 38.1% of variance ($R^2$) for each of total WMH, pWMH and dWMH, respectively.

| Region | | Hemisphere | $P_{FDR}$ | OR (95% CI) | $R^2$-gain |
|---|---|---|---|---|---|
| Precentral gyrus | Area 4, upper limb region | Contralesional | 0.038 | 8.45 (1.85-46.13) | 1.9% |
| | Caudal dorsolateral area 6 | Contralesional | 0.038 | 9.49 (2.02-54.44) | 2.5% |
| Middle frontal gyrus | Ventral area 9/46 | Contralesional | 0.038 | 8.84 (1.87-51.29) | 1.9% |
| | | Ipsilesional | 0.011 | 50.66 (5.07-1013.33) | 9.9% |
| | Area 46 | Ipsilesional | 0.011 | 50.66 (5.07-1013.33) | 9.9% |
| | Ventrolateral area 8 | Ipsilesional | 0.041 | 10.27 (1.80-79.17) | 1.8% |
| | Inferior frontal junction | Ipsilesional | 0.041 | 10.27 (1.80-79.17) | 1.8% |
| | Lateral area 10 | Ipsilesional | 0.038 | 13.47 (2.48-104.21) | 6.5% |
| Inferior frontal gyrus | Ventral area 44 | Ipsilesional | 0.041 | 10.27 (1.80-79.17) | 1.8% |
| | Inferior frontal sulcus | Contralesional | 0.038 | 10.50 (2.20-62.21) | 3.7% |
| | | Ipsilesional | 0.011 | 50.66 (5.07-1013.33) | 9.9% |
| Insular gyrus | Dorsal dysgranular insula | Ipsilesional | 0.041 | 10.27 (1.80-79.17) | 1.8% |
| Amygdala | Medial amygdala | Contralesional | 0.038 | 8.44 (1.90-44.24) | 2.3% |
| Basal ganglia | Globus pallidus | Contralesional | 0.038 | 8.45 (1.86-45.05) | 1.6% |
| Hippocampus | Rostral hippocampus | Contralesional | 0.038 | 8.45 (1.86-45.05) | 1.6% |
| Thalamus | Lateral pre-frontal thalamus | Contralesional | 0.038 | 9.73 (2.11-53.81) | 2.6% |
| | Pre-motor thalamus | Contralesional | 0.038 | 9.73 (2.11-53.81) | 2.6% |

**Supplementary Table 5. Sensitivity analysis where model was adjusted for pWMH-mediated global network damage instead of WMH-volume.**
Sensitivity analysis for significant associations between dichotomized pWMH-related network damage affecting cortical and subcortical brain regions and outcome after stroke, where model was adjusted for pWMH-mediated global network damage (i.e. mean ChaCo within the extended motor network) instead of WMH-volume. Thus, results are adjusted for age, initial NIHSS, lesion and pWMH-mediated global network damage. Model details are given in Supplementary Table 4. ORs with 95% CIs are given for patients with higher disconnectivity (reference) of rising one level in mRS compared to patients with lower disconnectivity for the specific region. *P* values are FDR corrected ($P_{FDR}$) for 93 tests. $R^2$-gain is given as additional explained variance ($R^2$) compared to $R^2$ of the modified base model (45.4%).

| Region | | Hemisphere | *P* | OR (95% CI) | $R^2$-gain |
|---|---|---|---|---|---|
| Precentral gyrus | Area 4 (upper limb region) | Contralesional | 0.001 | 7.25 (1.53-41.53) | 6.5% |
| | Caudal dorsolateral area 6 | Contralesional | 0.001 | 7.73 (1.51-49.21) | 5.7% |
| Middle frontal gyrus | Ventral area 9/46 | Ipsilesional | 0.001 | 32.13 (3.61-517.02) | 12.8% |
| | | Contralesional | 0.053 | 4.38 (0.98-22.21) | 3.7% |
| | Area 46 | Ipsilesional | 0.001 | 32.13 (3.61-517.02) | 12.8% |
| | Ventrolateral area 8 | Ipsilesional | 0.034 | 6.21 (1.15-39.69) | 3.4% |
| | Inferior frontal junction | Ipsilesional | 0.034 | 6.21 (1.15-39.69) | 3.4% |
| | Lateral area 10 | Ipsilesional | 0.014 | 7.37 (1.49-44.80) | 7.2% |
| Inferior frontal gyrus | Ventral area 44 | Ipsilesional | 0.034 | 6.21 (1.15-39.69) | 3.4% |
| | Inferior frontal sulcus | Ipsilesional | 0.001 | 32.13 (3.61-517.02) | 12.8% |
| | | Contralesional | 0.011 | 7.74 (1.57-46.76) | 7.5% |
| Insular gyrus | Dorsal dysgranular insula | Ipsilesional | 0.050 | 5.33 (1.00-33.19) | 3.1% |
| Amygdala | Medial amygdala | Contralesional | 0.047 | 4.63 (1.02-23.22) | 2.9% |
| Basal ganglia | Globus pallidus | Contralesional | 0.038 | 5.11 (1.09-27.02) | 2.9% |
| Hippocampus | Rostral hippocampus | Contralesional | 0.038 | 5.11 (1.09-27.02) | 2.9% |
| Thalamus | Lateral pre-frontal thalamus | Contralesional | 0.008 | 10.86 (1.83-89.11) | 8.0% |
| | Pre-motor thalamus | Contralesional | 0.008 | 10.86 (1.83-89.11) | 8.0% |

**Supplementary Table 6. Sensitivity analysis where patients with imagery not taken immediately after stroke onset were excluded.**

Sensitivity analysis for significant associations between dichotomized pWMH-related network damage affecting cortical and subcortical brain regions and outcome after stroke, where patients with imagery not obtained immediately after stroke onset (n=3) were excluded. Results are adjusted for age, initial NIHSS, lesion and total WMH volumes. ORs with 95% CIs are given for patients with higher disconnectivity (reference) of rising one level in mRS compared to patients with lower disconnectivity for the specific region. $R^2$-gain is given as additional explained variance ($R^2$) compared to $R^2$ of the base model (39.5%).

**Supplementary Figures**

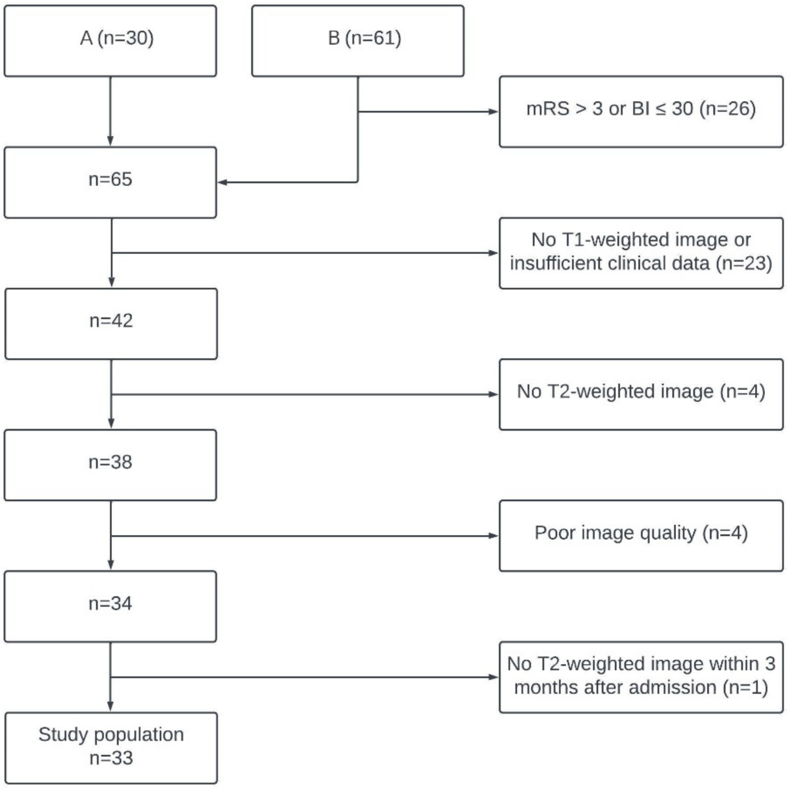


**Supplementary Figure 1. Dataset composition.**

Flowchart depicting inclusion and exclusion of participants, which are derived from 2 independent cohorts A[17] and B.[18]

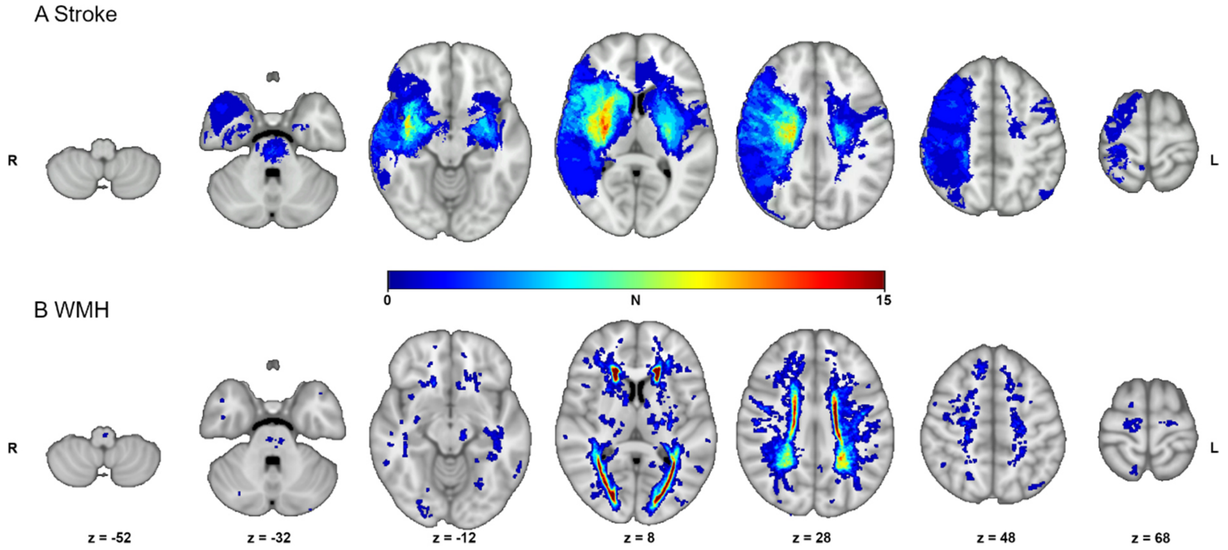


**Supplementary Figure 2. Distribution of stroke and WMH lesions.**
Lesion distribution heatmap for stroke lesions (A) and WMH (B), superimposed on a T1-weighted brain image in MNI standard space. The color represents the number (N) of patients having lesions within an area. The letters "R" and "L" indicate the side of the brain. Z-values indicate the horizontal slice in MNI standard space.

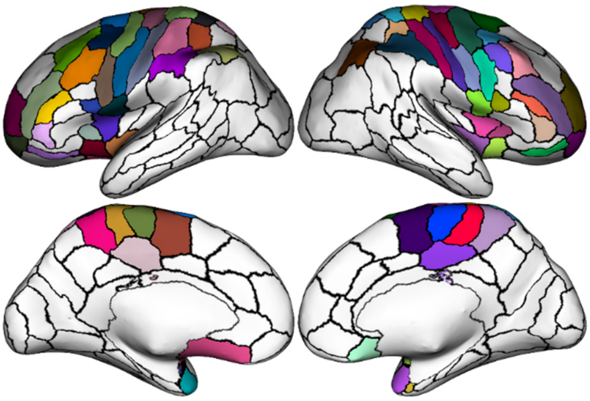

**Supplementary Figure 3. Extended Motor Network (cortical regions only).**
Cortical regions of the extended motor network, visualized as a brain surface overlay.
